# Effectiveness of Relaxation Therapy for Wound Healing in Patients with Diabetic Foot Ulcers: A Systematic Review of Randomized Control Trials

**DOI:** 10.1101/2025.04.26.25326493

**Authors:** Daanyal Noor Farrukh, Misbah Munaf Gaurd, Mohammad Dawood Farrukh, Fiza Farrukh-Hassan

## Abstract

Diabetic foot ulcers (DFU) is a serious complication of diabetes that leads to open sores found on the lower extremity, ultimately decreasing the quality of life. The purpose of this study is to systematically review randomized control trials (RCTs) to evaluate the effectiveness of relaxation therapy on healing wounds in patients with diabetic foot ulcers. The search was conducted across the databases of PubMed, Embase (Ovid), Cochrane Library, Google Scholar, and Web of Science. Inclusion criteria consisted of RCTs that compared relaxation therapy to standard care or other psychological/psychosocial intervention. The primary outcomes looked at were DFU healing, DFU extent, and perceived stress scale. The study included 3 RCTs that enrolled 75 patients with diabetic foot ulcers. It was found that within-intervention, relaxation therapy led to improved DFU healing, DFU extent, and participants were less stressed. Despite this, no evidence was found that suggested that relaxation therapy significantly improves outcomes compared to standard care or neutral imagery. Overall, it cannot be concluded that relaxation therapy has an effect on outcomes as there isn’t enough RCTs to come to a conclusion. Future research should delve into long-term effects of relaxation therapy on wound healing in patients with DFU, as well as compare it to other implemented therapies, such as cognitive behavioral therapy.

## 1.0 Introduction

Approximately 18.6 million people worldwide (including 1.6 million in the USA) are affected by diabetic foot ulcer (DFU) each year, often contributing to 80% of lower extremity amputations among diabetic patients (Armstrong et al., 2023). In the USA alone, the mean cost of treatment per patient per year who suffers from DFU can range from $3368 to $30131 (Lo et al., 2021). The two facts alone tell us that treatment for DFU can cost anywhere from 5.4 billion USD to 48 billion USD, putting significant burden on the healthcare cost (Armstrong et al., 2023; Lo et al., 2021). This financial strain is further compounded by the clinical challenges of managing DFUs. The most common prognosis of DFU is minor or major amputation, with their free survival rate showing 56.9% and 91.0% at 5 years, suggesting DFU is often slow to heal and highly recurrent (Armstrong et al., 2023). In addition to physiological factors such as peripheral arterial disease and diabetic neuropathy, psychological stress has emerged as a critical yet underrecognized barrier to wound healing (Wang et al., 2022; Fereira et al., 2023b). Chronic stress can impair immune function, elevate inflammatory responses, and disrupt tissue regeneration, factors that are especially detrimental in individuals with diabetes (Pradhan et al., 2009; Gouin et al., 2010; Kuebler et al., 2013).

While conventional management of DFUs involves glycemic control, debridement, pressure offloading, and infection prevention, recent attention has shifted toward the influence of psychosocial factors, particularly chronic psychological stress, in delaying wound healing (Wang et al., 2022; Fereira et al., 2023a; Fereira et al., 2023b; Pereira et al., 2025). Chronic stress is known to impair immune function, increase cortisol levels, prolong inflammation, and hinder angiogenesis, all processes critical to effective tissue repair.

Relaxation therapies, such as progressive muscle relaxation, and guided imagery, are potential tools to reduce stress in patients with diabetic foot ulcers. There have been many studies that synthesized the evidence to show that relaxation therapy is effective at treating different types of wounds, such as patients with burns, or patients who underwent surgery and are recovering (Kutenai et al., 2023; Broadbent et al., 2012). Despite this, there have been no prior systematic reviews that have comprehensively synthesized the evidence on relaxation therapy specifically as a therapeutic for DFU healing outcomes.

The objective of this systematic review is to evaluate the effectiveness of relaxation therapy on wound healing outcomes in patients with diabetic foot ulcers. Specifically, we aimed to assess changes in wound healing rates, DFU extent, and perceived stress levels through synthesis of literature that implements relaxation-based interventions alongside standard practices and compare it to a passive or active control group. In addition, we hope to assess the psychological benefits of relaxation therapy, such as reductions in stress, and how relaxation therapy affects pain levels, quality of life, and patient satisfaction. Overall, we will explore the feasibility and acceptability of incorporating relaxation techniques as adjunct therapies in DFU management protocols.

## 2.0 Methodology

### 2.1 INFORMATION SOURCES

A comprehensive search of the literature for primary sources was undertaken in the following sources: PubMed Central (PubMed), Embase (Ovid), Web of Science, Cochrane Library, and Google Scholar.

### 2.2 SEARCH STRATEGY

Each source was searched independently. The search strategy was developed using keywords and subject headings, where available, related to the diabetic foot ulcers AND wound healing AND relaxation therapy. The complete search strings for each database used are included in Supplementary S1. The article searches were limited to within the last 10 years (2015 – 2025) and english.

The search strategy for each source was peer-reviewed by two independent reviewers.

A final search was run, and references were exported on 17 April, 2025.

### 2.3 ELIGIBILITY CRITERIA

We established specific criteria for study inclusion in this systematic review. Studies of any design that reported on relaxation therapy and diabetic foot wounds were considered eligible. Clinical trials, including randomized clinical trials, involving human subjects were included. References were uploaded into Microsoft Excel for deduplication and screening. Two independent reviewers screened the abstracts of the identified studies and reviewed the full texts of the studies that were deemed potentially eligible. Disagreements between the reviewers were resolved through consensus discussions.

### 2.4 DATA COLLECTION AND EXTRACTION PROCESS

Data related to the application of relaxation therapy in the context of chronic wound healing were systematically extracted from each included study.

## 3.0 Results

Figure 1. Illustrates a flow diagram of our search results. It illustrated that initially 43 results were retrieved from 5 sources. After duplicates were removed, screening was narrowed down to 24 articles. After reviewing full-text articles, 7 studies were excluded due to being irrelevant, leaving 17 full-text articles for screening. Further screening resulted in 13 articles being excluded for not meeting inclusion/exclusion criteria. The excluded articles included articles that utilized wrong outcome of measures, or were not randomized control trials. Table 1. Illustrates key information for all included studies.

**Table 1.** Description of the studies investigating the effects of different relaxation therapies in participants with diabetic foot ulcers. I/C: Intervention/Control; RCT: Randomized Control Trial; DM: Diabetes Mellitus; DFU: Diabetic Foot Ulcers; PSS: Perceived Stress Scale; EG: Experimental Group; ACG: Active Control Group; PCG: Passive Control Group.

| First author/<br>Year/<br>Country | Study Design | Study Duration | Sample I/C | Population | Intervention | Control | Outcome Measures | Results |
| --- | --- | --- | --- | --- | --- | --- | --- | --- |
| Ferreira et al. (2023a), Portugal | Qualitative Study Nested in an RCT | 8 weeks | 8/0 | Adults ($\geq 18$ years) with DM and chronic DFUs lasting $>6$ weeks. | Four individual 45-minutes sessions of muscle relaxation and positive imagery. | None | DFU Healing Score, PSS, perspectives from patients. | Improved DFU healing, decreased PSS, greater adherence to treatments (e.g. |
|  |  |  |  |  |  |  |  | foot off-loading), |
| Ferreira et al. (2023b), Portugal | Three-Arm RCT | 8 weeks; 6 month follow-up | 54 participants randomized: EG (n=21), ACG (n=17), PCG (n=16) | Adults aged ≥18 years with DM and 1–2 active chronic DFUs lasting >6 weeks. |  | ACG (Neutrel Imagery Session); PCG (No Treatment) | DFU Healing Score, DFU Extent, Perceived Stress Scale | EG/ACG showed significant decrease in PSS, DFU extent and an improvement in DFU healing. |
| Pereira et al. (2025), Portugal | Longitudinal Pilot RCT | 8 weeks | 60 participants randomized into 1 of 4 groups using a 1:1:1:1 ratio. | 18+ with diabetes and 1-2 chronic DFUs lasting >6 weeks but <14 weeks who visited a diabetic foot outpatient clinic in Portugal. | Progressive muscle relaxation with guided imagery receiving a 45-minute sessions every 2 week | Standard medical treatments for DFUs with no psychological intervention. | DFU Healing, PSS | Improved DFU Healing and PSS within-treatment groups. |

**Figure 1.**
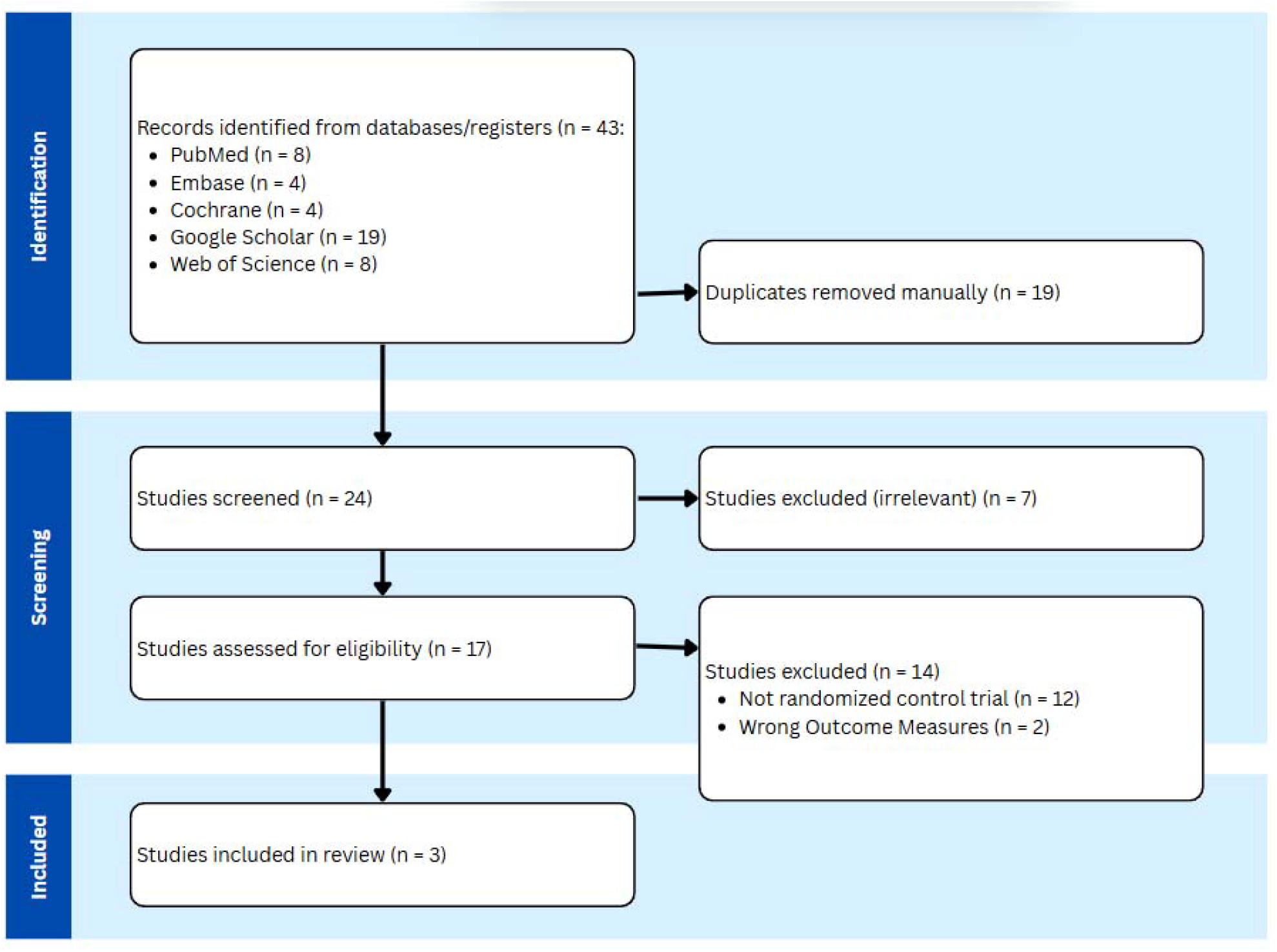
PRISMA Flow Diagram; This diagram shows the systematic process we followed to include papers captured by our search.

### 3.1 DIABETIC FOOT ULCER HEALING AND EXTENT

Two articles evaluated wound healing progress across interventions using the Resvech 2.0 assessment tool, whereas one article looked at DFU extent as well (Pereira et al., 2025; Ferreira et al., 2023a).

In a research study by Pereira et al. (2025), DFU healing was measured using the Resvech 2.0 assessment tool (Pereira et al., 2025). Significant improvements within intervention groups were noted between the baseline and post-intervention for patients that were administered muscle relaxation therapy (β = −5.88, p < 0.001) as well as between the baseline and follow-up (β = −8.39, p < 0.001) (Pereira et al., 2025). This information is supported in a study by Fereira et al. (2023a), where it was found that DFU healing score had significantly increased from baseline to post-intervention, and baseline to follow-up (β = −5.67, p < 0.001; β = −8.20, p < 0.001). Furthermore, there were significant differences between the intervention and passive control group in DFU healing (β = 4.54, p = 0.02) (Pereira et al., 2025). Despite this, Fereira et al. (2023a) contradicts the findings of Pereira et al. (2025), indicating that DFU healing is not significantly different between intervention and passive control groups (β = 2.816,p > 0.05).

In a research study by Fereira et al. (2023a), significant improvements were noted in DFU extent within the intervention group between the baseline and post-intervention (β = −3.45, p < 0.05) as well as between the baseline and follow-up (β = −3.55, p < 0.05).

### 3.2 PERCEIVED STRESS SCALE

Two articles evaluated perceived stress scale (PSS) across interventions (Pereira et al., 2025; Ferreira et al., 2023a).

In a research study by Pereira et al. (2025), significant improvements within intervention groups were noted between the baseline and post-intervention for patients that were administered muscle relaxation therapy (β = −5.25, p < 0.005), and between the baseline and follow-up (β = −4.26, p > 0.05). Furthermore, it was found that PSS had significantly increased from baseline to post-intervention, and baseline to follow-up (β = −5.38, p < 0.001; β = −4.61, p< 0.05) (Fereira et al., 2023a). However, there were no significant differences between post-intervention and passive control groups in PSS (β = 1.59, p > 0.05) (Pereira et al., 2025).Fereira et al. (2023a) reinforces the findings of Pereira et al. (2025), indicating that PSS is not significantly different between intervention and passive control groups (β = 2.647, p > 0.05).

### 3.3 QUALITATIVE MEASURES

In Fereira et al. (2023b), five themes emerged that addressed the suitability of relaxation therapy for the treatment of diabetic foot ulcers: perception of intervention, perception regarding distress, perception regarding technique, changes in the patient’s life, and changes in DFU.

Patients with DFUs reported significant emotional distress, particularly fear of amputation. Witnessing others with amputations heightened anxiety, and some expressed hopelessness even after surgery, highlighting the psychological burden associated with the condition.

Patients were unfamiliar with relaxation techniques but reported feeling calm, relieved, and emotionally uplifted after sessions. Some noted physical benefits, including improved circulation and foot sensitivity. Overall, participants were highly satisfied and recommended the intervention to others with DFUs.

The relaxation intervention promoted more positive thinking among patients, shifting focus away from fear and toward healing. Some also reported improved interpersonal relationships, highlighting broader emotional and social benefits beyond physical symptom relief.

Patients noticed improvements in their DFUs, including reduced wound size, but expressed uncertainty about relaxation’s direct role in healing. Many viewed it as a complementary approach that eased psychological stress and potentially supported the body’s healing process.

## 4.0 Discussion

Overall, the systematic review found that relaxation therapy, including progressive muscle relaxation and guided imagery, was associated with significant within-group improvements in DFU healing, DFU extent, and PSS from baseline to follow-up (Pereira et al., 2025; Fereira et al., 2023a). This suggests that relaxation therapy is an effective treatment for DFU, as it improves healing, decreases DFU extent and results in patients having lower outcomes of distress and stress long-term (Pereira et al., 2025; Fereira et al., 2023a). Despite this, when compared to an active control group that received 4 neutral imagery sessions, it was found that there was no significant difference between DFU healing, DFU extent, and PSS (p > 0.05), suggesting that relaxation therapy, though effective at healing, does not produce any different outcomes from other psychosocial treatments. To place these findings in a broader context, it is important to consider prior literature evaluating the role of psychological interventions and stress in wound healing.

Comparing findings to other studies on psychological interventions in chronic wound healing or diabetes management, we find that psychological and psychosocial interventions yield positive effects on wound healing (Robinson et al., 2017; Muhrawi et al., 2022). In a systematic review, Robinson et al. (2017) highlighted that psychological interventions consistently yield a positive outcome on wound healing, reporting effect sizes ranging from 0.53 to 1.89 across surgical wounds. Muhrawi et al. (2022) identified this correlation as a critical risk factor in individuals with chronic wounds, reinforcing the notion that addressing psychological well-being is paramount in managing these patients. This suggests the need for more empathy-centered care, as validating concerns can improve patient outcomes before surgery and postoperative during the wound healing process.

Moreover, psychological factors have been shown to establish a relationship with diabetic foot ulcers, where mental health can both impede and reflect the state of wound healing. More broadly, the relationship between stress and wound healing is directly addressed by the treatment of relaxation therapy for stress in patients (Fereira et al., 2023b). Similar interventions that target stress reduction have been associated with healing delays, indicating that psychosocial factors can influence outcomes (Robinson et al., 2017; Gouin & Kiecolt-Glaser, 2012). These findings align with meta-analytic evidence suggesting a statistically significant negative correlation between stress and wound healing across various studies (Walburn et al., 2009; Gouin & Kiecolt-Glaser, 2012).

The interplay between stress and wound healing is noteworthy due to multiple biological mechanisms that mediate this relationship as research has shown that psychological stress can significantly hinder wound healing by altering hormonal responses, inflammatory pathways, and immune system functions. Glucocorticoids play a major role in this process, as elevated cortisol levels occur due to activation of the hypothalamic-pituitary-adrenal axis while under stress (Herman et al., 2016). Increased glucocorticoid levels inhibit the expression of cytokines, such as interleukin-1, which are crucial pro-inflammatory cytokines that initiate the wound healing process (Marucha et al., 1998; Glaser et al., 1999). Furthermore, deficiencies in keratinocyte growth can be disrupted by stress as it disrupts glucocorticoid secretion, decreasing the extent of skin repair (Marucha et al., 1998).

In addition, delayed healing can be caused by a heightened level of catecholamine due to stress, such as epinephrine and norepinephrine, leading to vasoconstriction and a reduction in blood flow to the wound, shrinking the oxygen supply and nutrient delivery to repair mechanisms (Romana-Souza et al., 2010; Razjouyan et al., 2017; Jozic et al., 2017). This overall results in a significantly impaired healing process (Muhrawi et al., 2022; Pradhan et al., 2009).

The role of the immune system is also pivotal, as studies have indicated that psychological stress correlates with diminished expression of inflammatory mediators such as interleukin-8 and macrophage inflammatory protein-1α (Pradhan et al., 2009; Gouin et al., 2010). This is further reinforced by Kuebler et al. (2013), where it was found that stress is linked to a reduction in immune response, as macrophages exhibit impaired microbicidal activity in the presence of stress hormones. Overall, stress has been linked to affect both early and late inflammatory phases of wound healing, which is crucial for preventing infections and promoting adequate healing respectively (Christian et al., 2006; Glaser et al., 1999).

Stress adversely affects wound healing through a multifaceted mechanism involving hormonal changes, altered immune responses, and disruptions in inflammatory signaling pathways, which collectively undermine the body’s natural repair processes.

Clinically, these findings suggest that integrating stress-reduction techniques into standard DFU care protocols may not only benefit wound healing outcomes, but also improve patient engagement, emotional well-being, and adherence to recommended treatments such as foot offloading (Fereira et al., 2023b). Given the relatively low cost and non-invasive nature of relaxation therapies, their use as complementary interventions warrants broader clinical investigation and implementation.

Despite this, the systematic review is limited due to a number of variables. This review utilized a small number of included studies, resulting in the review not being conclusive. On top of this, all articles included took place in Portugal, further limiting the generalizability of the therapy on the greater population. Additionally, the intervention and control groups revealed mixed results, failing to indicate that relaxation therapy is superior to other psychosocial interventions. The studies failed to effectively blind participants and personnel as psychologists employed to carry out the treatment were not blinded to what treatment they would be delivering.

In the future, more research should delve into the possibility of implementing this therapy complementary to other interventions in diverse settings with standardized protocols. Research that includes biomarkers of stress and long-term ulcer recurrence should be conducted to come to a more conclusive answer. Relaxation therapy should also be compared to other more widely accepted psychological/psychosocial therapies, such as cognitive behavioural therapy, guided imagery, and standard education. If clinical implementation becomes a goal in the future, researchers need to conduct an economical analysis of this treatment to measure the cost-effectiveness of relaxation therapy.

## 5.0 Conclusion

This systematic review evaluated the impact of relaxation therapy on wound healing outcomes among patients with diabetic foot ulcers (DFUs). The findings suggest that relaxation therapies, such as progressive muscle relaxation and guided imagery, are associated with significant within-group improvements in DFU healing, extent, and perceived stress levels. However, when compared to active or passive controls, no significant differences were observed, indicating that relaxation therapy may be beneficial but not necessarily superior to other psychosocial interventions. Importantly, the review highlights the potential role of psychological stress as a modifiable factor in wound healing, consistent with broader literature linking stress reduction to improved physiological outcomes. Nevertheless, the limited number of high-quality randomized controlled trials, small sample sizes, and lack of geographic diversity restrict the generalizability of the conclusions. Moreover, methodological limitations, including insufficient blinding, further temper the strength of the evidence. Future research should focus on larger, multicenter trials that directly compare relaxation therapy to other psychosocial interventions, incorporate objective biomarkers of stress, and assess long-term clinical outcomes, including ulcer recurrence and quality of life. Additionally, economic evaluations are warranted to determine the cost-effectiveness of integrating relaxation therapies into standard DFU care. Overall, while preliminary evidence is promising, more rigorous research is needed to substantiate the clinical utility of relaxation therapy as an adjunctive treatment for diabetic foot ulcer management.

## Data Availability

All data produced in the present work are contained in the manuscript.

## Notes

### Competing Interest Statement

The authors have declared no competing interest.

### Funding Statement

This study did not receive any funding

