## Supplemental Table 1 S1 for "Effectiveness of Relaxation Therapy for Wound Healing in Patients with Diabetic Foot Ulcers: A Systematic Review of Randomized Control Trials"

**Supplementary Table S1. Systematic Review Search Terms.**

| Database | Search Strategy |
| --- | --- |
| PubMed | ("Diabetic Foot Ulcer"[tiab] OR "Diabetic Foot Ulcers"[MeSH Terms] OR "DFU"[tiab] OR "Chronic Diabetic Wound"[tiab] OR "Diabetic Wound"[tiab]) AND ("Wound Healing"[MeSH Terms] OR "Ulcer Healing"[tiab] OR "Chronic Wound"[tiab]) AND ("Relaxation Therapy"[MeSH Terms] OR "Relaxation"[tiab] OR "Relaxation Training"[tiab] OR "Stress Reduction"[tiab] OR "Guided Imagery"[tiab] OR "Hypnosis"[MeSH Terms] OR "Hypnosis"[tiab] OR "Hypnotherapy"[tiab] OR "Self-Hypnosis"[tiab]) |
| Embase (Ovid) | 1. exp diabetic foot ulcer/ or "diabetic foot ulcer".ti,ab. or DFU.ti,ab. or "diabetic wound".ti,ab.  2. exp wound healing/ or "wound healing".ti,ab. or "ulcer healing".ti,ab. or "chronic wound".ti,ab.  3. exp relaxation therapy/ or exp relaxation/ or relaxation.ti,ab. or "relaxation training".ti,ab. or "stress reduction".ti,ab. or "guided imagery".ti,ab. or "progressive muscle relaxation".ti,ab. or "mind body intervention".ti,ab. or "breathing exercise".ti,ab. or "autogenic training".ti,ab.  4. 1 and 2 and 3  5. limit 4 to (randomized controlled trial)  6. 5 not (conference abstract or editorial or letter).pt. |
| Web of Science | "TS=(""diabetic foot ulcer"" OR DFU OR ""diabetic wound"") AND TS=(""wound healing"" OR ""ulcer healing"" OR ""chronic wound"") AND TS=(""relaxation therapy"" OR ""relaxation training"" OR ""progressive muscle relaxation"" OR ""stress reduction"" OR ""guided imagery"" OR ""autogenic training"" OR ""breathing exercises"" OR ""mind-body intervention"") Clinical Trial or Article, 2015-2025" |
| Google Scholar | "diabetic foot ulcer" AND "wound healing" AND ("relaxation therapy" OR "guided imagery" OR "breathing exercises") AND ("randomized trial" OR "clinical trial") |
| Cochrane Library | ("diabetic foot ulcer" OR "diabetic wound" OR DFU) AND ("wound healing" OR "ulcer healing" OR "chronic wound") AND ("relaxation" OR "relaxation therapy" OR "relaxation training" OR "stress reduction" OR "progressive muscle relaxation" OR "guided imagery" OR "autogenic training" OR "breathing exercises" OR "mind-body intervention") |
